# Inability to work following COVID-19 vaccination among healthcare workers - an important aspect for future booster vaccinations

**DOI:** 10.1101/2022.11.21.22282594

**Authors:** Julia Reusch, Isabell Wagenhäuser, Alexander Gabel, Anna Höhn, Thiên-Trí Lâm, Lukas B. Krone, Anna Frey, Alexandra Schubert-Unkmeir, Lars Dölken, Stefan Frantz, Oliver Kurzai, Ulrich Vogel, Manuel Krone, Nils Petri

## Abstract

**Background:** COVID-19 vaccination is a key prevention strategy to reduce the spread and severity of SARS-CoV-2 infections, especially among highly exposed healthcare workers (HCWs). However, vaccine-related inability to work among HCWs could overstrain healthcare systems.

**Methods:** This study examined sick leave and intake of pro re nata (PRN) medication after the first, second and third COVID-19 vaccination in HCWs. Subgroup analyses were performed for different vaccines, gender, healthcare professions, and for HCWs aged at least 30 years. Data was collected by using an electronic questionnaire.

**Findings:** Among 1,704 HCWs enrolled, in total 595 (34·9%) HCWs were on sick leave following at least one COVID-19 vaccination, leading to a total number of 1,550 sick days. Both the absolute sick days and the rate of HCWs on sick leave significantly increased with each subsequent vaccination. Comparing BNT162b2mRNA and mRNA-1273 the difference in sick leave was not significant after the second dose, but mRNA-1273 induced a significantly longer and more frequent sick leave after the third.

**Interpretation:** A considerable number of HCWs have been on sick leave after COVID-19 vaccination, staff absences increase with each additional dose, depend on the vaccine, and vary between HCWs’ gender, and profession. In the light of further COVID-19 infection waves and booster vaccinations, there is a risk of additional staff shortages due to post-vaccination inability to work, which could acutely overload healthcare systems and jeopardise patient care. These findings will aid further vaccination campaigns to minimise the impact of staff absences on the healthcare system.

**Funding:** This study was funded by the Federal Ministry for Education and Science (BMBF) via a grant provided to the University Hospital of Wuerzburg by the Network University Medicine on COVID-19 (B-FAST, grant-No 01KX2021) as well as by the Free State of Bavaria with COVID-research funds provided to the University of Wuerzburg, Germany. Nils Petri is supported by the German Research Foundation (DFG) funded scholarship UNION CVD.

## 1 Introduction

The ongoing COVID-19 pandemic,^1^ with its high morbidity and accompanying therapeutic demands, has drastically demonstrated the urgency of a resilient public health system.^2^ Healthcare workers (HCWs) of all qualifications play a central role in the critical infrastructure needed to cope with the high SARS-CoV-2 infection prevalence levels.^3-6^

The COVID-19 pandemic has a direct impact on the availability of public healthcare due to the persistent and non-negligible burden of disease. The exceptional strain on health systems is caused by the increasing number of patients but also by the number of HCWs on sick leave due to a SARS-CoV-2 infection.^7^

COVID-19 vaccination has emerged as a key strategy to control the spread and severity of SARS-CoV-2 infections, particularly among HCWs. However, vaccine-related incapacity could overwhelm public healthcare and must be addressed as part of this important prevention strategy. Vaccine-related staff absences must be considered in light of future COVID-19 booster vaccination campaigns and the challenges posed by the continuing COVID-19 pandemic.^8^ To date evidence is insufficient to predict the impact of COVID-19 vaccine-related incapacity on healthcare systems.^9-11^

The main goal of this study was to examine the number of sick days as well as the rate of staff absences after the first, second and third dose of COVID-19 vaccination among HCWs. We also performed a detailed analysis to assess the intake of pro re nata (PRN) medication following vaccinations and examined the impact of vaccine type, gender, age, and profession on the inability to work following COVID-19 vaccination.

### Research in context

**Evidence before this study**

To assess the evidence published to date, PubMed and medRxiv were searched for the terms „sick days“, „sick leave“, inability to work”, “healthcare workers” in combination with „COVID-19 vaccination“, „COVID-19 vaccine“, „SARS-CoV-2 vaccine” in the title or abstract published between the 1^st^ of January 2020 and the 30^th^ of October 2022.

The evidence to date on inability to work associated with the administration of a COVID-19 vaccine, particularly with regard to HCWs as key public stakeholders and in the setting of a real-life point-of-care investigation, is still insufficient. There is a lack of data on a cohort covering a broad age spectrum. Especially the impact of a third ‘booster’ dose of COVID-19 vaccine on the inability to work is still unclear.

Previous studies have described increased staff absences after administration of mRNA-1273 compared to BNT162n2mRNA, as well as in younger and female HCWs. All studies published to date have examined only basic immunisation, including administration of the first and second COVID-19 vaccine dose.

**Added value of this study**

This study presents the first large-scale, real-world analysis of sickness-related absence following COVID-19 vaccination in HCWs, particularly after administration of a third vaccine dose. The data shows that the rate of sick leaves increased with the number of vaccinations. In the comparison of mRNA-based vaccines, mRNA-1273 caused higher rates of inability to work than BNT162b2mRNA.

**Implications of the available evidence**

Particularly in view of further COVID-19 booster vaccinations in the context of emerging virus variants of concerns (VOC) and upcoming SARS-CoV-2 infection peaks, sick days following COVID-19 vaccination need to be taken into account. The selected cohort of HCWs is highly exposed and absences of HCWs will have a direct impact on public health capacity. Our results will inform the design of vaccination campaigns and provide a rational for optimisation measures such as the staggering of vaccination offers.

## 2 Methods

### 2.1 Study setting

The study presented was conducted as part of the prospective CoVacSer cohort study, which examines the progression of Anti-SARS-CoV-2-Spike IgG levels as well as quality of life and work ability in HCWs after COVID-19 vaccination and/or SARS-CoV-2 infection.

Participants who met all of the following inclusion criteria were recruited for the CoVacSer study: (i) age ≥ 18 years, (ii) written informed consent, (iii) minimum interval of 14 days after first confirmation of SARS-CoV-2 infection by polymerase chain reaction (PCR) and/or at least one dose of COVID-19 vaccination regardless of vaccination schedule, (iv) employment in healthcare sector. Vaccination schedules with vaccines that were not licensed by the European Medicines Agency (EMA) during the data collection period were excluded from the data analysis according to the study protocol.^12^

Under the study protocol, participants were followed up at set intervals for 24 months after the last SARS-CoV-2 immunising event, with the follow-up cycle restarting with each new SARS-CoV-2 infection or COVID-19 vaccination. Serum blood samples in combination with pseudonymised digital questionnaires were collected both at study enrolment and at each follow-up.

Since the 18^th^ of November 2021, COVID-19 booster vaccinations have been officially offered at the earliest six months after the second dose of COVID-19 vaccine or after the last SARS-CoV-2 infection, according to recommendation of the German Standing Committee on Vaccination (STIKO). For immunocompetent adults aged 18-30 years, a single dose of BNT162b2mRNA (30µg) was recommended; for those aged ≥30 years, a single dose of BNT162b2mRNA (30µg) or half a dose of mRNA-1273 (50μg). Both heterogeneous and homologous vaccine combinations were recommended.^13^

### 2.2 Data collection

The data collection period ranged from the 29^th^ of September 2021 to the 27^th^ of March 2022.

Only participants who had been vaccinated at least once against COVID-19 without convalescence of SARS-CoV-2 before their first vaccination were included into data analysis. All further follow-up examinations were incorporated as long as participants were not yet infected with SARS-CoV-2.

For the first vaccination, the COVID-19 vaccines BNT162b2mRNA, mRNA-1273, ChAdOx1-S and Ad26.CoV2-S were considered. For the second and third vaccination, only participants who had received BNT162b2mRNA and/or mRNA-1273 were included, in accordance with the recommendations of the German Standing Committee on Vaccination (STIKO) during the data collection period.^13^

Mainly HCWs from a single tertiary hospital were involved in this study, but HCWs from surrounding hospitals and medical surgeries were also included.

The questionnaire including questions on sick leave and post-vaccination medication needs was conducted using the platform REDCap (Research Electronic Data Capture, projectredcap.org).^14^

Occupational groups were asked in the following sections: “nursing”, “physicians”, “other HCWs with patient contact” and “other HCWs without any patient contact”.

### 2.3 Ethical approval

The study protocol was approved by the Ethics committee of the University of Wuerzburg in accordance with the Declaration of Helsinki (file no. 79/21).

### 2.4 Statistics

The statistical analyses were performed with the statistical programming language R (version 4.2.1). For determining statistical differences, such as sick leave and intake of PRN medication, within pairwise comparisons of HCWs’ subpopulations, exact Fisher tests were performed. For comparing the days of sick leave between HCWs grouped by COVID-19 vaccination doses, gender, and profession, Wilcoxon rank-sum tests were performed. Mean days of sick leave after vaccinations were presented as bar graphs with error bars indicating one standard deviation. The percentage of HCWs were also presented as bar graphs, whereas the error bars indicate the upper confidence interval based on a binomial distribution. To account for multiple testing, p-values were corrected based on Benjamini-Yekutieli.^15^

## 3 Results

### 3.1. Participant recruitment and characterisation of the study population

From the 29^th^ of September 2021 to the 27^th^ of March 2022, 1,831 individuals participated and completed the study questionnaire.

1,704 (93·1%) individuals were eventually included in the data analysis. 77 participants did not meet the study inclusion criteria according to the study protocol, 42 were convalescent of SARS-CoV-2 infection prior to their first COVID-19 vaccination and eight had received a COVID-19 vaccine other than the ones considered. A detailed overview of excluded participants is provided in *Figure 1*.

**Figure 1.**
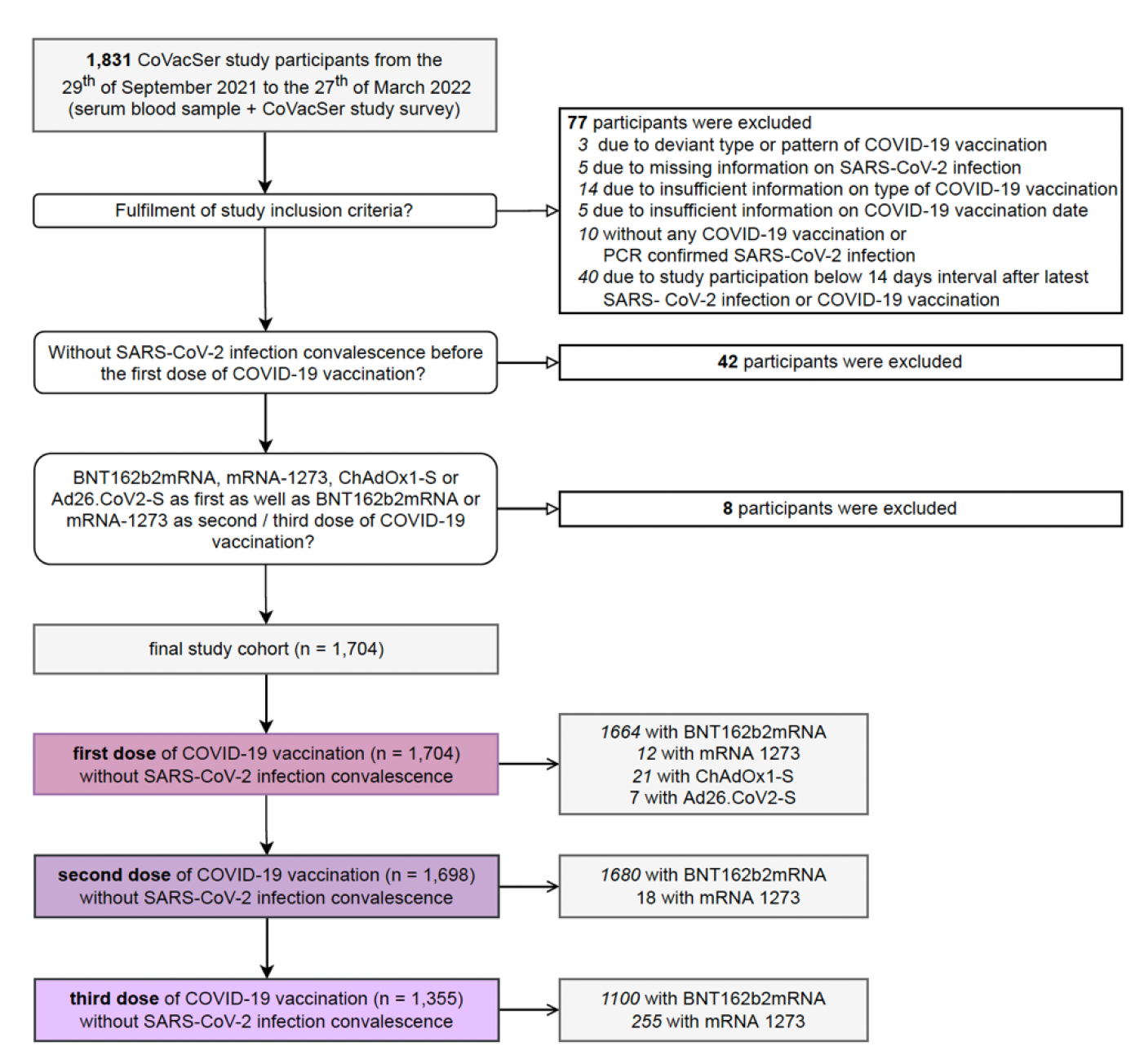
Recruitment of study participants and characterisation of final study population separated by number of doses and type of COVID-19 vaccine

81·0% (1,379/1,704) of the included participants identified as female, 19·0% (325/1,704) as male, with an overall median age of 39 (IQR: 29-52) years.

Overall, all 1,704 included participants received at least a first dose of COVID-19 vaccine, 99·6% (1,698/1,704) a second and 79·5% (1,355/1,704) a third one, without any SARS-CoV-2 infection prior to the respective vaccine administration. 97·7% (1,664/1,704) were vaccinated with BNT162b2mRNA for the first administration, 0·7% (12/1,704) with mRNA-1273, 1·2% (21/1,704) with ChAdOx1-S and 0·4% (7/1,704) with Ad26.CoV2-S. BNT162b2mRNA accounts for 98·9% (1,680/1,698) of the second COVID-19 inoculations, while mRNA-1273 constitutes 1·1% (18/1,698). In the third COVID-19 vaccination dose, BNT162b2mRNA amounts to 81·0% (1,100/1,355), mRNA-1273 to 19·0% (255/1,355; *Figure 1*).

The enrolled HCWs were distributed among the four occupational groups as follows: 34·9% (594/1,704) nursing, 18·6% (317/1,704) physicians, 22·0% (375/1,704) other HCWs with patient contact and 24·5% (417/1,704) other HCWs without any patient contact. A detailed characterisation by profession is presented in *Table 1*.

**Table 1.**
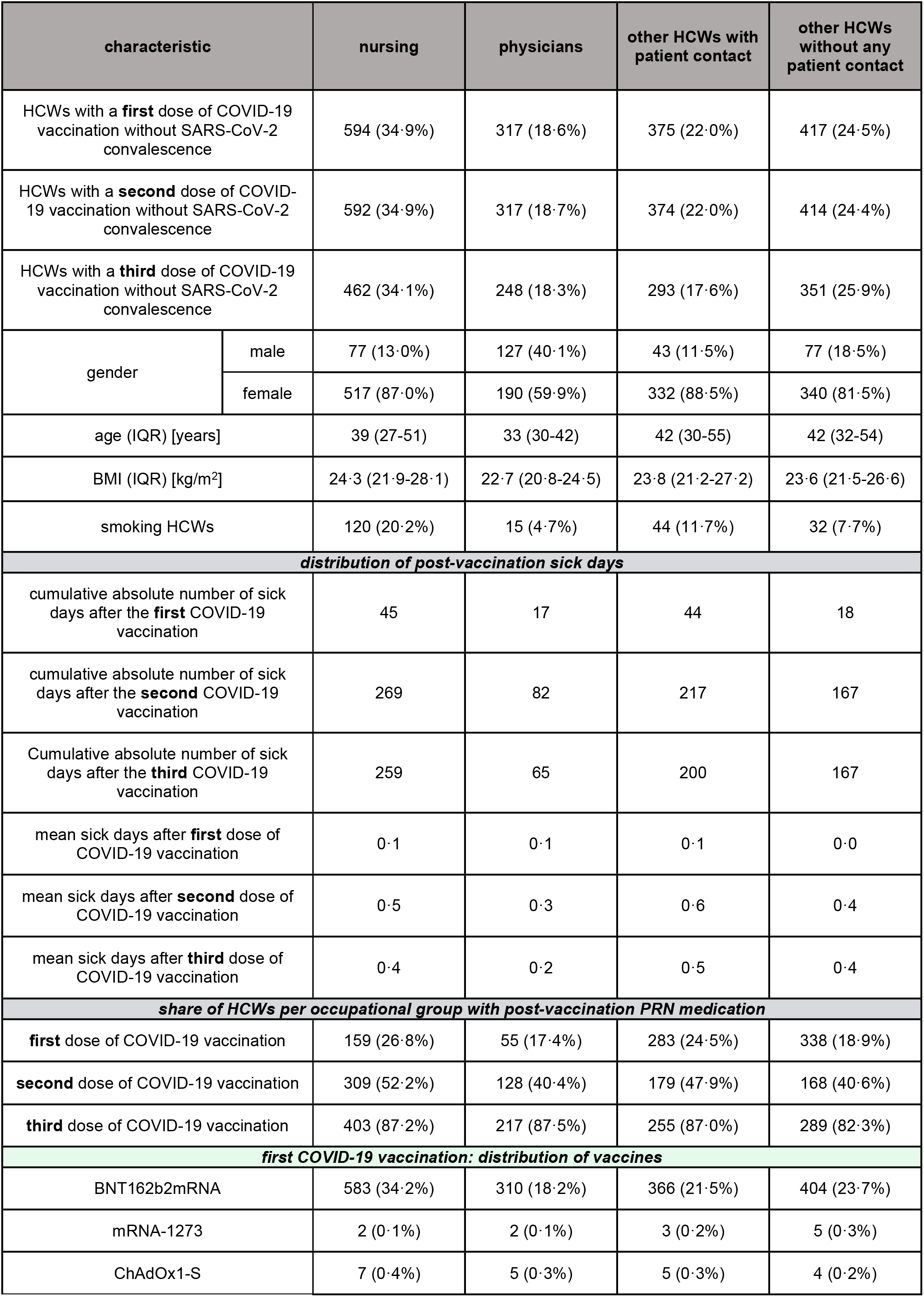
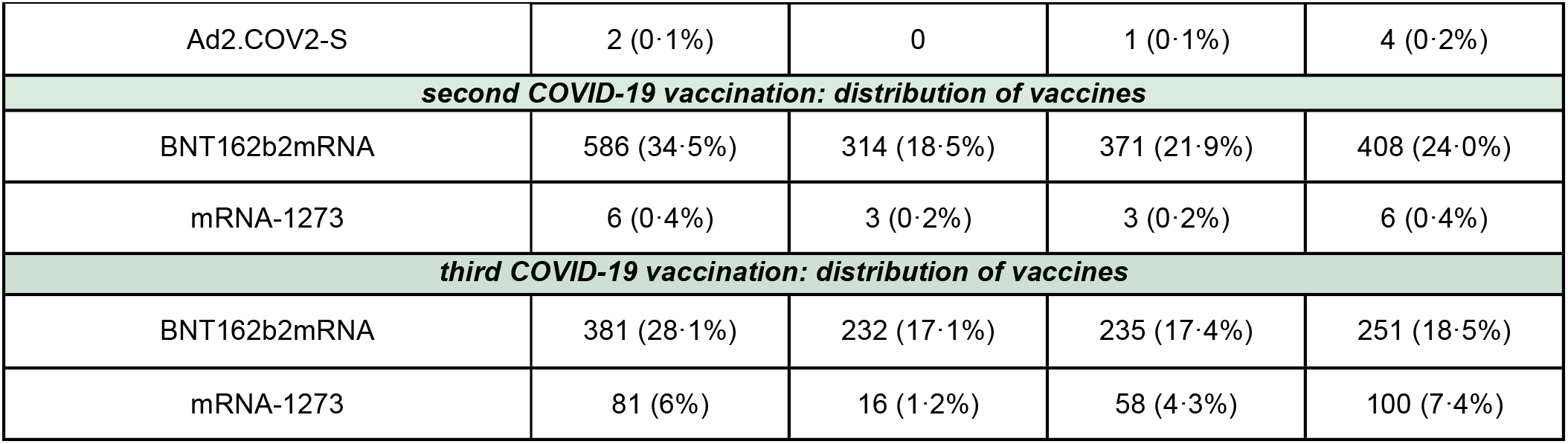
Characterisation of the study population separated by profession, percentages indicate the share of the study cohort. Age and BMI are reported as medians with interquartile ranges in brackets.

### 3.2 Sick leave separated by number of doses of COVID-19 vaccination

In total, absenteeism due to vaccine-related illness resulted in an absolute number of 1,550 sick days in the cohort of 1,704 participants for the entirety of COVID-19 vaccinations administered.

#### 3.2.1 Sick leave after the first dose of COVID-19 vaccination among HCWs

After the first COVID-19 vaccination dose, a total of 3·8% (51/1,704) HCWs were on sick leave, 3·1% (51/1,664) of those vaccinated with BNT162b2mRNA (0·1 mean sick days, 95 absolute sick days), none in case of mRNA-1273 administration, 52·4% (11/21) among ChAdOx1-S administered HCWs (1·1 mean sick days, 22 absolute sick days) and 28·6% (2/7) of those vaccinated with Ad26.CoV2-S (1·0 mean sick days, 7 absolute sick days).

The proportion of HWCs who were absent from work after a first dose of ChAdOx1-S was significantly higher than after administration of BNT162b2mRNA (p<0·0001) or mRNA-1273 (p<0·05; *Figure 2A*). Mean sick days after a first dose of ChAdOx1-S were significantly more than after administration of BNT162b2mRNA (p<0·0001) or mRNA-1273 (p<0·05); mean sick days after a first dose of Ad26.CoV2-S were significantly longer than after administration of BNT162b2mRNA (p<0·001; *Figure 2D*).

**Figure 2.**
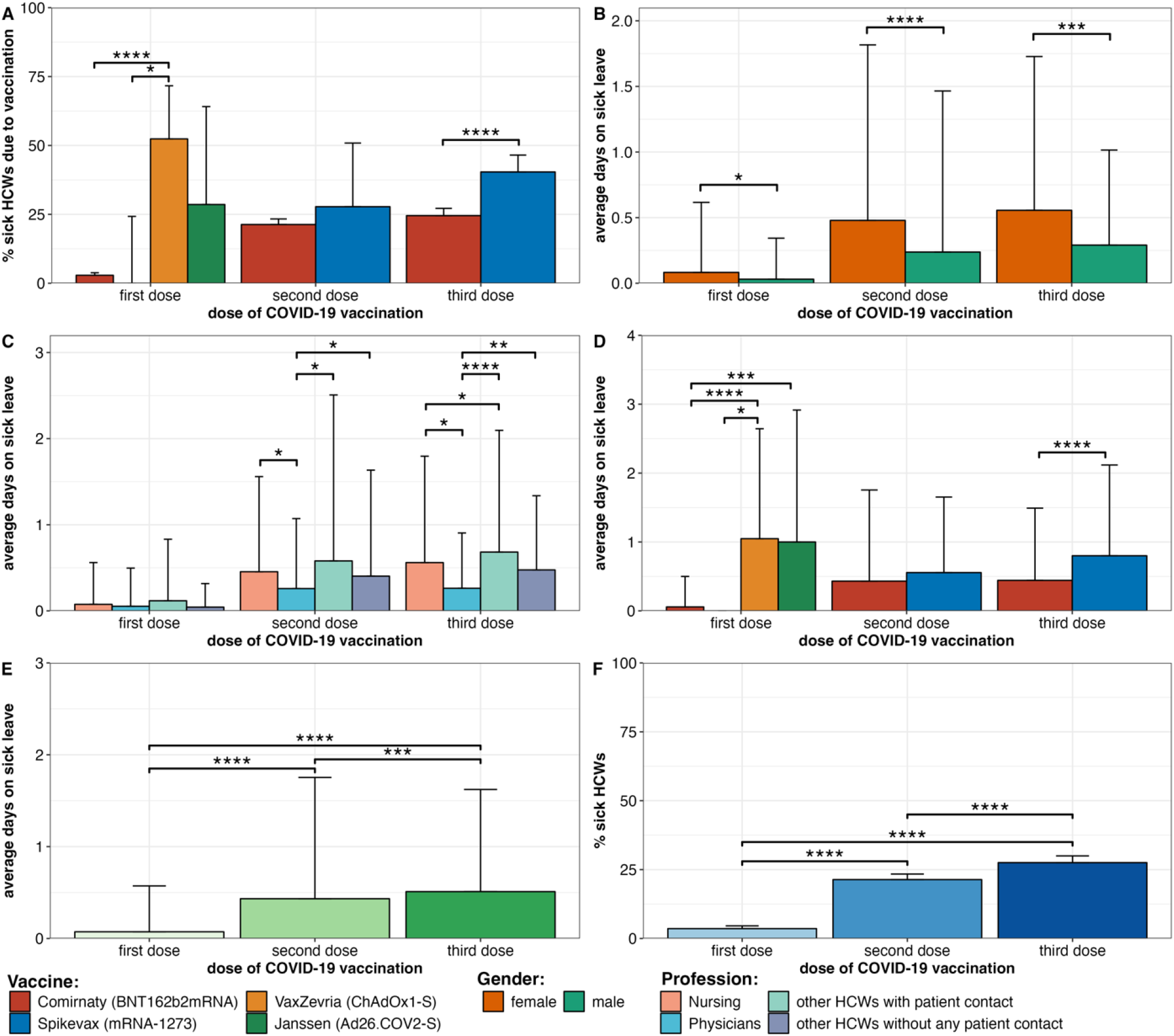
Comparison of sick leave depending on the number of doses and type of COVID-19 vaccine. Figure **2A** portrays the relative proportion of HCWs on sick leave stratified by number of doses and type of COVID-19 vaccine. Figure **2B** pictures the average days on sick leave stratified by number of doses and gender, **2C** by number of doses and profession **2D** by number of doses and type of COVID-19 vaccine. **2E** visualises the overall average sick leave days stratified by number of doses and **2F** the relative share of HCWs on sick leave separated by number of doses. **2A&F:** Error bars indicate the corresponding confidence interval, **2B-E:** Bar height and error bars indicate group means and one standard deviation. *: p < 0·05 **: p < 0·01 ***: p < 0·001 ****: p < 0·0001

#### 3.2.2 Sick leave after the second dose of COVID-19 vaccination among HCWs

In the case of the second COVID-19 vaccination, 21·8% (370/1,698) of HWCs reported incapacity for work. 21·7% (365/1680) within the group of HCWs vaccinated with BNT162b2mRNA (0·4 mean sick days, 725 absolute sick days) and 27·8% (5/18) among HCWs administered mRNA-1273 (0·6 mean sick days, 10 absolute sick days).

After the second dose of COVID-19 vaccination, there was no significant difference in the proportion of HCWs on sick leave or mean sick days after administration of BNT162b2mRNA or mRNA-1273 (*Figure 2A and 2D*).

#### 3.2.3 Sick leave after the third dose of COVID-19 vaccination among HCWs

Following the third dose of COVID-19 vaccination, overall, 27·9% (378/1,355) of HCWs were off sick, 24·7% (272/1100) of those vaccinated with BNT162b2mRNA (0·4 mean sick days, 487 absolute sick days) and 41·6% (106/255) of participants that received mRNA-1273 (0·8 mean sick days, 204 absolute sick days).

After a third COVID-19 vaccination dose, significantly more HCWs were on sick leave after mRNA-1273 compared to BNT162b2mRNA administration (p<0·0001; *Figure 2A*) and mean sick days after mRNA-1273 were significantly more than after BNT162b2mRNA administration (p<0·0001; *Figure 2A and 2D*).

#### 3.2.4 Post-vaccination sick leave among HCWs, separated by gender

Separated by gender, 4·1% female HCWs (56/1,379) were on sick leave after the first COVID-19 vaccination, 23·7% (325/1,374) following the second and 29·7% (234/1,121) after the third vaccine administration. In case of male HCWs, 1·5% (5/325) were on sick leave after the first vaccination, 11·7% (38/324) after the second and 17·1% (40/194) after the third one (*Figure 2B)*.

Overall, 526 of 1,379 (38·1%) female and 69 of 325 (21·2%) male HCWs were on sick leave, *s*howing a significant ratio of absent female compared to male HCWs (p<0·0001). Separated by dose of COVID-19 vaccinations for all three administrations female HCWs showed significantly higher mean sick days (p<0·05 for the first, p<0·0001 for the second, p<0·001 for the third one; *Figure 2B*)

#### 3.2.5 Post-vaccination sick leave among HCWs, separated by occupational groups

Separated by occupational groups, among nursing HCWs 3·4% were on sick leave following the first COVID-19 vaccination (0·1 mean sick days, 45 absolute sick days), 22·3% after the second (0·5 mean sick days, 269 absolute sick days) and 25·6% after the third one (0·4 mean sick days, 259 absolute sick days).

Among physicians, 2·8% were on sick leave after the first (0·05 mean sick days, 17 absolute sick days), 15·5% after the second (0·3 mean sick days, 82 absolute sick days) and 17·3% after the third vaccination (0·2 mean sick days, 65 absolute sick days).

Among other HCWs with patient contact, 5·3% were off sick after the first vaccination (0·1 mean sick days, 44 absolute sick days), 24·3% after the second (0·6 mean sick days, 217 absolute sick days) and 36·9% after the third one (0·5 mean sick days, 200 absolute sick days).

Among HCWs without any patient contact, 2·9% were on sick leave following the first vaccination (0·0 mean sick days, 18 absolute sick days), 22·7% after the second (0·4 mean sick days, 167 absolute sick days) and 29·6% after the third administration (0·4 mean sick days, 167 absolute sick days; *Figure 2D*).

At the second COVID-19 vaccination, the mean number of sick days was significantly higher among nursing HCWs compared to physicians (p<0·05); physicians were significantly less likely to be on sick leave than other HCWs with patient contact (p<0·05) and then other HCWs without any patient contact (p<0·05; *Figure 2C*).

Following the third COVID-19 vaccination, nurses recorded significantly longer mean sick leave compared to physicians (p<0·05), but fewer mean sick days than other HCWs with patient contact (p<0·05); physicians had significantly shorter sick leave compared to other HCWs with (p<0·0001) and without patient contact (p<0·01; *Figure 2C)*.

#### 3.2.6 Overall sick leave after the first, second and third dose of COVID-19 vaccination among HCWs

Overall, 3·8% (51/1,704) of HCWs were on sick leave (0·1 average sick days) after the first dose of COVID-19 vaccination, 21·8% (370/1,698; 0·4 average sick days) after the second and 27·9% (378/1,355; 0·5 average sick days) after the third one. With each additional dose, there was a significant increase in the proportion of HCWs on sick leave (p<0·0001, comparing first and second dose; p<0·0001, comparing first and third dose; p<0·0001, comparing second and third dose; *Figure 2F)* as well as in the mean sick leave (p<0·0001, comparing first and second dose; p<0·0001, comparing first and third dose; p<0·001, comparing second and third dose; *Figure 2E*).

### 3.3 Comparison of BNT162b2mRNA and mRNA-1273 as booster vaccines in HCWs aged ≥30 years

Since mRNA-1273 was officially recommended as a booster vaccine in Germany only for individuals at least 30 years of age,^13^ an age-adjusted subgroup analysis of the third COVID-19 vaccination considering HCWs only aged ≥30 years was conducted (*Table 2*).

**Table 2.** Characterisation of the study population aged ≥30 years at the time of the third COVID-19 vaccination; percentages indicate the share of the sub-cohort HCWs ≥30 years, separated by vaccination. Age and BMI are reported as medians with interquartile ranges in brackets.

| HCWs with a third dose of COVID-19 vaccine, aged $\geq 30$ years | | BNT162b2mRNA | mRNA-1273 |
| --- | --- | --- | --- |
| absolute number of study participants |  | 806 (76%) | 252 (24%) |
| gender | male | 159 (19.7%) | 34 (13.5%) |
|  | female | 647 (80.3%) | 218 (86.5%) |
| age (IQR) [years] |  | 46 (37-55) | 46 (39-55) |
| BMI (IQR) [kg/m<sup>2</sup>] |  | 24.2 (21.8-27.5) | 23.8 (21.5-27.1) |
| smoking [absolute number] |  | 102 (12.7%) | 24 (9.5%) |
| cumulative number of post-vaccination **sick days** after COVID-19 booster vaccination |  | 341 | 204 |
| **number of HCWs** on post-vaccination sick leave after COVID-19 booster vaccination |  | 193 (24.0%) | 106 (42.1%) |
| HCWs with post-vaccination **PRN medication** after COVID-19 booster vaccination |  | 75 (9.3%) | 71 (28.2%) |
| **distribution to occupational groups** |  |  |  |
| nursing |  | 258 (24.4%) | 80 (7.6%) |
| physicians |  | 175 (16.6%) | 15 (1.4%) |
| other HCWs with patient contact |  | 175 (16.6%) | 58 (5.5%) |
| other HCWs without any patient contact |  | 197 (18.6%) | 99 (9.4%) |

Within this subgroup of HCWs aged ≥30 years, after the third COVID-19 vaccination, 23·7% (191/806) inoculated with BNT162b2mRNA were off sick (0·4 mean sick days, 341 absolute sick days) compared to 40·9% (103/252) of HCWs vaccinated with mRNA-1273 (0·8 mean sick days, 204 absolute sick days).

Thus, mRNA-1273 administration was responsible for a significantly higher proportion of HCWs on sick leave than BNT162b2mRNA (p<0·0001; *Figure 3A*); also, the mean number of sick days was significantly higher among mRNA-1273 booster vaccinees (0·8 mean sick days, 202 absolute sick days) than among HCWs administered with BNT162b2mRNA (0·3 mean sick days, 902 absolute sick days) (p<0·0001, *Figure 3B*).

**Figure 3.**
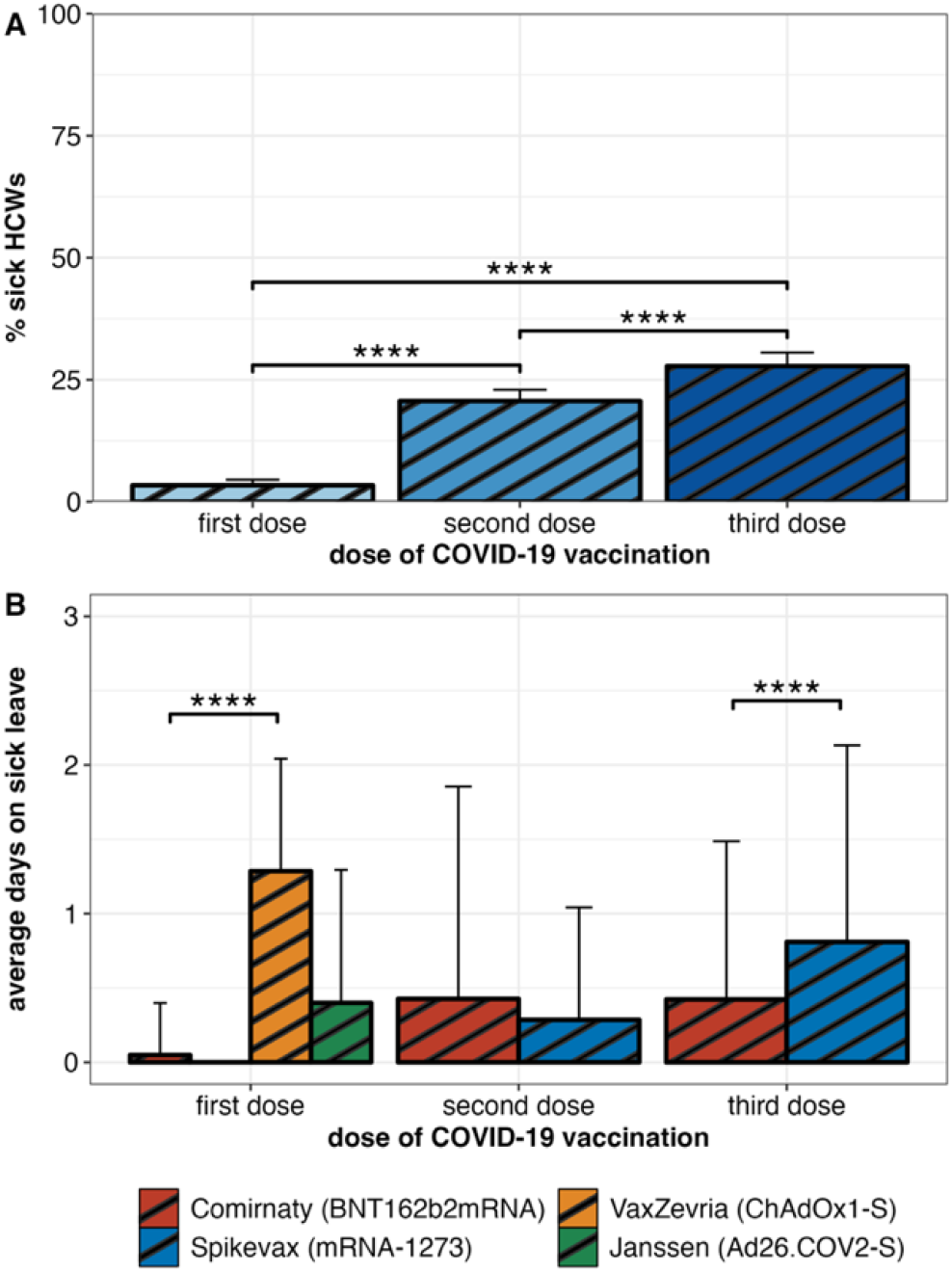
Comparison of sick leave depending on the number of doses of COVID-19 vaccine, adjusted for HCWs ≥30 years. Figure **3A** portrays the relative number of HCWs on sick leave administered with mRNA-1273, **3B** the average days on sick leave separated by COVID-19 vaccine, both stratified by number of doses of COVID-19 vaccine. Bar height and error bars indicate group means and one standard deviation. ****: p < 0·0001

### 3.4 Post-vaccination sick leave in relation to PRN medication

After administration of the first COVID-19 vaccine dose, a total of 22·6% (385/1,704) HCWs reported intake of PRN medication, after the second dose 46·2% (784/1,698) and after the third dose 86·0% (1,165/1,355; *Figure 4A)*.

**Figure 4.**
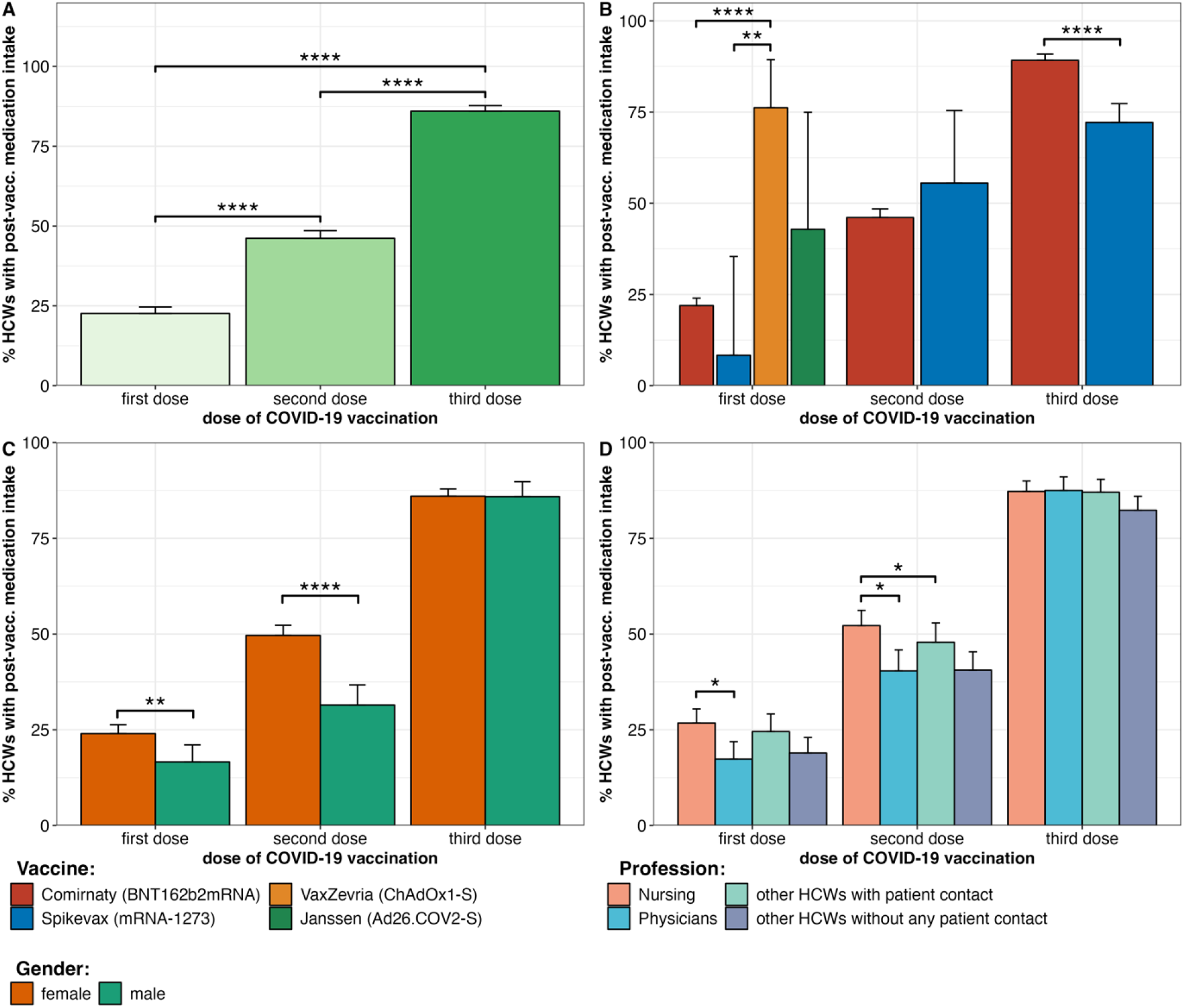
Post-vaccination PRN medication, separated by the number of doses, type of COVID-19 vaccine, gender, and profession **Figure 4A** portrays the percentage of HCWs with PRN medication after COVID-19 vaccine administration, separated by the number of doses, **4B** separated by the number of doses and type of vaccine, **4C** separated by number of doses and gender, and **4D** separated by the number of doses and profession. Bar height and error bars indicate group means and upper border of confidence interval. *: p < 0·05 **: p < 0·01 ****: p < 0·0001

The proportion of HCWs using PRN medication after the first vaccine dose was 21·9% (365/1,668) for BNT162b2mRNA, 8·3% (1/12) for mRNA-1273, 76·2% (16/21) for ChAdOx1-S and 42·9% (3/7) for Ad26.CoV2-S.

Following the second COVID-19 vaccination, 46·1% (774/1,680) of HCWs vaccinated with BNT162b3mRNA required PRN medication, 55·6% (10/18) in case of mRNA-1273.

After the third dose, the proportion of HCWs using PRN medication was 89·2% (981/1,100) for BNT162b2mRNA-vaccinated and 72·2% (184/255) for mRNA-1273-vaccinated.

After the first vaccine dose, HCWs reported a significantly higher rate of medication use after administration of ChAdOx1-S compared to both BNT162b2mRNA (p<0·0001) and mRNA-1273 (p<0·01). After the third dose, a significantly higher percentage of HCWs vaccinated with BNT162b2mRNA reported the use of PRN medication compared to HCWs vaccinated with mRNA1273 (p<0·0001; *Figure 4B*).

Separated by gender, 24·0% (331/1,379) of female and 16·6% (54/325) of male HCWs took PRN medication after the first COVID-19 vaccination, 49·6% (682/1,374) female and 31·5% (102/324) male HCWs after the second vaccine administration, and 86·0% (964/1,121) female and 85·9% (201/234) male HCWs after the third one.

Female HCWs had a significantly higher proportion of post-vaccination medication use after the first (p<0·01) and second (p<0·0001) administration of COVID-19 vaccination compared to male HCWs. No significant difference was found between female and male HCWs for the third COVID-19 vaccination *(Figure 4C)*.

By occupation, 26·8% (159/594) of nurses, 17·4% (55/317) of physicians, 24·5% (283/375) of other HCWs with and 18·9% (338/417) of HCWs without any patient contact indicated the use of PRN medication following the first COVID-19 vaccination. For the second COVID-19 vaccination, 52·2% (309/592) of nurses, 40·4% (128/317) of physicians, 47·9% (179/374) of other HCWs with and 40·6% (168/414) of HCWs without any patient contact reported the use of PRN medication. Regarding the third COVID-19 vaccination, 87·2% (403/462) of nurses, 87·5% (217/248) of physicians, 87·0% (255/293) of other HCWs with and 82·3% (289/351) of HCWs without patient contact used PRN medication (*Table 1*).

A significantly higher share of enrolled nurses compared to physicians reported taking medication PRN after the first (p<0·05) and second (p<0·05) administration of COVID-19 vaccine, as well as in comparison to the group of other HCWs with patient contact after the second vaccination (p<0·05; *Figure 4D)*.

## 4 Discussion

Overall, COVID-19 vaccination caused a relevant number of sick days among HCWs. HCWs who were administered the mRNA-based vaccines BNT162b2mRNA and mRNA-1273 as their first dose of COVID-19 vaccination had little post-vaccination sick leave. In contrast, a higher number of HCWs that were administered ChAdOx1-S or Ad26.CoV2-S as their first COVID-19 vaccination were subsequently on sick leave. The first dose of ChAdOx1-S caused a significantly higher rate of sick leave compared to the mRNA-based vaccines and we observed a similar, albeit not statistically significant, result in the small group (n=7) of HCWs receiving Ad26.CoV2-S. Comparing BNT162b2mRNA and mRNA-1273, the administration of mRNA-1273 resulted in a significantly higher rate of post-vaccination sick leave after the third vaccination dose, with a similar trend for the second vaccination dose. The higher rate of sick leave might be explained by differences in vaccine composition. Following mRNA-1273 vaccination, increased anti-poly(ethylene) glycol (PEG) antibodies have been associated with increased reactogenicity.^16^ A comparatively higher rate of PRN medication intake after BNT162b2mRNA vaccination could have reduced the rate of sick leave. Previous studies report that the immunogenicity of mRNA-1273 with its proportionally higher dosage exceeds that of BNT162b2mRNA and leads to a more pronounced vaccination response.^17,18^ Our finding that mRNA-1273 causes more sick leave supports these results and is very robust as the higher rate of sick leave is significant in the overall analysis and in the more homogenous subgroup analysis of enrolled HCWs aged ≥30 years.

In contrast to the first COVID-19 vaccine dose, the second dose resulted in an increase both in the rate of HCWs on sick leave and in the average duration of sick leave, which further intensified after the third dose respectively.

The extent of sick leave following subsequent doses of COVID-19 vaccination can be expressed in the rate of cumulative sick days per 1,000 HCWs. In our representative cohort, we found 74·5 sick days per 1,000 HCWs for the first vaccination dose, 437·5 for the second dose, and 510·0 for the third dose. Consequently, COVID-19 vaccination has a non-negligible impact on the staff availability in the health sector. Albeit that COVID-19 vaccination is undeniably an important infection prophylaxis for HCWs, which aims to avoid quarantine- and infection-related staff shortages, the vaccination also causes a temporary relevant impairment of staff availability due to sickness absences. The economic leverage of these sickness-related absences, due to limited treatment capacities of hospitals, should be further considered.^19-21^

The observed rise in sick leave with increasing repetitions of COVID-19 vaccinations might be explained by an immune response boost that is amplified with each subsequent vaccination. This could lead to an augmentation of vaccine-related reactions causing side effects and incapacity for work, which is in line with previous findings of increasing humoral and T-cellular immunogenicity as well as with reports about more vaccine-related side effects with repeated vaccinations.^8,22^ Comparison of different occupational groups among the HCWs revealed that physicians spent fewer days on sick leave after the second and third vaccination than other professions. In addition, the lowest rate of post-vaccination drug intake was reported among physicians. In comparison, the rates of sick leave as well as PRN medication intake were higher among nurses. The increased use of medication might be an indicator of more severe side effects and hence inability to work. This difference might be explained by demographic variables such as age, gender and health status (e.g. BMI). In addition, the different work routine of the nursing and medical staff, which entails a higher physical burden for the nursing staff, might further explain the higher sickness rate among nurses. Furthermore, significantly fewer male than female HCWs used PRN medication after the first and second COVID-19 vaccination.

Our study has some relevant limitations. Since HCWs were selected as study population, the collective is predominantly female, with women accounting for 81·0% of all participants, reflecting the typical socio-demographic composition of HCWs in Germany.^23^ Data on side effects and sick leave were collected retrospectively, so a potential influence of recall bias and subjectivity cannot be ruled out. Dosage and time of PRN medication intake after vaccination was not queried. The number of sick days after vaccination was not evaluated separately for working days and weekends or holidays, which could reduce the extent of sick leave if HCWs were incapable on their days off. Due to varying vaccine availability and the recommendations of the German Standing Committee on Vaccination (STIKO),^13,24^ the frequencies of the different COVID-19 vaccines in our cohort are unequally distributed for the first, second and third vaccinations, respectively, however, this reflects a real-life scenario. Since the federal recommendation for administration of mRNA-1273 as a booster vaccine was restricted to individuals who were ≥30 years old,^13^ data on sick leave among HCWs younger than 30 years vaccinated with mRNA-1273 as booster vaccine are lacking. Of all the COVID-19 vaccines considered, BNT162b2mRNA was the most frequently administered vaccine. The number of HCWs vaccinated with ChAdOx1-S (n=21) and Ad26.CoV2-S (n=7) in our study was small and these two vaccines were only used for the first COVID-19 vaccination dose. As the focus of our study was on mRNA-based COVID-19 vaccines, our results on sick leave after administration of ChAdOx1-S and Ad26.CoV2-S should be interpreted with caution. Individuals immunised with the COVID-19 subunit vaccine NVX-CoV2373 were not included in this study as it was approved by the European Medicines Agency (EMA) after the study recruitment period and not recommended for a third vaccine dose in Germany in individuals without contraindication for the use of mRNA vaccines. Further, due to the study period, the meanwhile available Omicron VOC adapted, bivalent COVID-19 vaccines are not included in the study. However, all of these limitations mentioned are direct consequences of the real-life study setting, which underlines the transferability of the data and is a particular strength of our study.

In conclusion, inability to work following COVID-19 vaccination among HCWs is an important and not-negligible aspect of this vital infection prevention measure. Staff absences related to vaccination are considerably shorter compared to those after COVID-19 infection.^25^ Therefore, vaccine-related absence from work cannot be an argument for non-vaccination. However, vaccination-related staff absences can have a direct impact on healthcare capacity and places an additional burden on an already understaffed healthcare systems dealing with demographic change, the additional burden of COVID-19 patients and shortages of qualified staff, especially nurses. To ensure patient safety and avoid work overload, staff shortages due to SARS-CoV-2 infected HCWs must be avoided. Our study provides important data to estimate the burden of sick leave among HCWs following COVID-19 vaccination. Our results provide representative data that will inform the planning of vaccination campaigns and provides a rational for optimisation measures such as the staggering of vaccination offers. In view of the planned COVID-19 booster vaccinations with a fourth or fifth dose, our data on a rise of sick leave with repeated vaccine doses highlights the need to closely monitor vaccine-related incapacity to work during the early stages of the vaccination campaigns.

## Data Availability

Additional data that underlie the results reported in this article, after de-identification (text, tables, figures, and appendices) as well as the study protocol, statistical analysis plan, and analytic code is made available to researchers who provide a methodologically sound proposal to achieve aims in the approved proposal on request to the corresponding author.

## 5 Author contributions

All authors had unlimited access to all data. Julia Reusch, Isabell Wagenhäuser, Alexander Gabel, Manuel Krone, and Nils Petri take responsibility for the integrity of the data and the accuracy of the data analysis.

*Conception and design*: Thiên-Trí Lâm, Lukas B. Krone, Anna Frey, Alexandra Schubert-Unkmeir, Lars Dölken, Stefan Frantz, Oliver Kurzai, Ulrich Vogel, Manuel Krone, Nils Petri.

*Trial management*: Julia Reusch, Isabell Wagenhäuser, Anna Höhn, Manuel Krone, Nils Petri.

*Statistical analysis*: Julia Reusch, Isabell Wagenhäuser, Alexander Gabel.

*Obtained funding*: Oliver Kurzai, Ulrich Vogel.

*First draft of the manuscript*: Julia Reusch, Isabell Wagenhäuser, Alexander Gabel, Lukas B. Krone, Manuel Krone, Nils Petri.

The manuscript was reviewed and approved by all authors except Professor Ulrich Vogel. He had a major contribution to the concept and design of the study as well as in obtaining funding. As he passed away on the 4^th^ of October 2022 during manuscript drafting, he was not able to review and approve the final version of the manuscript. We miss him as an enthusiastic college and friend who showed a great dedication to his work, family, and friends.

## Abbreviations

PCR: 
HCWs: 
PRN: 
VOC: 

