## Supplementary Table 1 for "Inability to work following COVID-19 vaccination among healthcare workers - an important aspect for future booster vaccinations"

### Supplements

**Supplementary Table 1**: Characteristics of study population separated by gender

| characteristic | female HCWs | male HCWs |
|---|---|---|
| absolute number of subjects (share on the study population) | 1,379 (81%) | 325 (19%) |
| Age (IQR) [years] | 39 (29-53) | 37 (31-48) |
| BMI (IQR) [kg/m$^2$] | 23·4 (21·1-26·7) | 24·5 (22·6-27·1) |
| smoking [absolute number] (share among the gender group) | 163 (11·8%) | 48 (14·8%) |
| **vaccine-related sick leave** | | |
| number of sick days after the first COVID-19 vaccination | 114 | 10 |
| number of sick days after the second COVID-19 vaccination | 658 | 77 |
| number of sick days after the third COVID-19 vaccination | 623 | 68 |
| number on HCWs on sick leave after first COVID-19 vaccination (share among the gender group) | 59 (4·3%) | 5 (1·5%) |
| number on HCWs on sick leave after second COVID-19 vaccination (share among the gender group) | 331 (24%) | 39 (12%) |
| number on HCWs on sick leave after third COVID-19 vaccination (share among the gender group) | 338 (24·5%) | 40 (12·3%) |
| **post-vaccination PRN medication** | | |
| number of HCWs with post-vaccination PRN medication, first dose administration (share among the gender group) | 331 (24·0%) | 54 (16·6%) |
| number of HCWs with post-vaccination PRN medication, second dose administration (share among the gender group) | 682 (49·6%) | 102 (31·5%) |
| number of HCWs with post-vaccination PRN medication, third dose administration (share among the gender group) | 964 (86·0%) | 201 (85·9%) |

***Supplementary Table 1**: Characterisation of the study population separated by gender; percentages indicate the respective share of the female and male sub-cohort. Age and BMI are reported as medians with interquartile ranges in brackets.*
